# Effectiveness of a Pain Neuroscience Education program on the physical activity of patients with Chronic Low Back Pain compared to a standard back school program, a randomized controlled study - END-LC

**DOI:** 10.1101/2023.09.18.23295477

**Authors:** Guillaume Thébault, Claire Duflos, Gaël Le Perf

## Abstract

**Introduction:** Education is recognized as an effective and necessary method in chronic low back pain. Nevertheless, the data on the effectiveness of education in physical activity in the medium or long term are not yet well known, nor the factors that could lead to practice such or such education. Our study aims to measure the effectiveness of a pain neuroscience education compared to a back school on physical activity three months and one year after educational sessions coupled with a multidisciplinary rehabilitation program. The study also seeks to measure the effects of these two types of education on several other factors including the intensity of pain and psycho-behavioral factors. Finally, it aims to identify the determinants of the success of the educational sessions coupled with the rehabilitation program.

**Methods and analysis:** The study will involve eighty-two adults with chronic low back pain. The study will be monocentric, prospective, open, controlled, and randomized, of superiority, with two parallel arms with an experimental group “pain neuroscience education” and a control group “back school”. The primary outcome will be the average number of steps taken at home over a week measured by an actigraph. Secondary outcomes include behavioral assessments. Descriptive and inferential analysis will be carried out on the primary and secondary judgment criteria. Multivariate modeling will be carried out using actimetric data and data from the main and secondary outcomes.

**Ethics and dissemination:** a favorable opinion was given by the Committee for Personal Protection of Ile de France VII on June 22, 2023 (National number: 2023-A00346-39). The study was previously registered with the National Agency or the Safety of Medicines and Health Products (IDRCB : 2023-A00346-39). This protocol is the version submitted to the Committee for Personal Protection of Ile de France VII entitled “Protocol Version N°1 of 03/29/2023”.

**Trial Registration:** NCT05840302.

**Article Summary:** **Strengths and limitations of this study**

- The use of an actigraph as a measuring tool of physical activity in ecological context to evaluate effectiveness of an education in low back pain.
- The follow-up at three months and one year after the rehabilitation program.
- The study includes an analysis of actimetric data, behavioral, occupational and psychological variables to determine the predictive factors for the success of educational sessions.
- The monocentric design of this study is a limitation.

## INTRODUCTION

### Background and rationale

Low back pain continues to be a challenge for researchers, clinicians and all healthcare professionals. It is a challenge to understand and model multiple processes entangled in low back pain and more widely in the persistence of pain or symptoms, to improve primary care or rehabilitation and cognitive therapies [1–4]. Low back pain is a major health problem in terms of frequency in the global population and health costs despite theoretical advances, new approaches and numerous clinical studies [5]. Indeed, recent epidemiological data show the importance of the problem of chronic low back pain. They indicate a 50% increase in chronic low back pain over the past twenty years and represents the 6th leading cause in the world in terms of disability [6, 7].

Education and interdisciplinary rehabilitation are treatments that have shown evidence of efficacy. They are highlighted for people suffering from chronic low back pain in most international recommendations and international authors have recently emphasized their interest [1].

Firstly, education aims to change misconceptions that people with chronic low back pain may have due to prior beliefs, avoidant attitudes, or catastrophic thoughts that have become established over the course of life history of chronicization of pain.

Historically, the first education program in this field was developed in Sweden under the name of back school [8]. The basis of the education was the presentation of biomechanical aspects such as the increase in interdiscal pressure during physical stress. The presentation of ergonomic posture aimed to help patients “protect” their back, and then prevent future spinal pathologies. Subsequently, many variants developed in North American countries. For example, Penttinen and colleagues [9] proposed ten lessons to increase physical activity in daily life and to train participants to ergonomic work techniques. This education is well suited to the biomedical model since it emphasizes the biological or mechanistic character of back pain: the pain emanates from a mechanical dysfunction that persists in the spine. Two literature reviews showed weak or conflicting evidence for the effectiveness of back school, from very heterogeneous studies [10, 11]. However, when the back school is based on a biopsychosocial model in addition to a rehabilitation program, this is recommended in certain guidelines [12].

Indeed, these last decades’ changes in therapeutic approaches are developed from the biopsychosocial model with integration of psychological aspects and social factors [13]. As an extension of this paradigmatic change, other types of education have developed, such as pain neuroscience education [14]. It consists of didactic learning of the physiological mechanisms of pain, understanding the influence of psychoaffective factors and central neurological processes. In other words, the chronic low back pain patient is given an understanding of pain as emerging from the dynamics of multiple processes and not from a single stable mechanism. Fundamentally, chronic pain is embodied and alters perceptual processes [15, 16].

Moseley and colleagues [14,17] showed that pain neuroscience education to be more effective than school back education based on anatomy and biomechanics. In addition, reviews of the literature have shown that pain neuroscience education has positive effects in the short and medium terms on the perception of pain, disability, catastrophism and on the improvement of physical performance when it is coupled with a rehabilitation program based on physical exercises [18,19]. Nevertheless, further investigations are needed to know the effectiveness beyond six months [20].

Through this quick literature review, we observe that the two educational techniques (back school vs pain neuroscience education) differ in the fundamental approaches on which they are based (biomedical vs biopsychosocial). Debates on the conceptual approach to low back pain are still current [21, 22]. Beyond these questions, it is important to know what educational content has a positive and lasting influence on people with low back pain.

Secondly, rehabilitation can be considered as an adjunctive treatment option that focuses on physical activity in order to fight against disability [1]. In view of the importance of physical activity, in France, the message delivered by health insurance is: “good treatment is movement”^1^ because some mechanisms (e.g. fear-avoidance) leads to a significant reduction in physical activity with deleterious consequences for the person with low back pain (in terms of social, professional, family repercussions and the physiological consequences that this entails). In this line, a recent review showed that the level of physical activity was associated with the prevalence of chronic low back pain: people with a medium level of physical activity have a 10% lower risk of low back pain than people with low level [23]. Thus, the measurement of physical activity in daily life is of relevant interest in the context of chronic low back pain in order to assess the effect of a rehabilitation program and educational sessions and to measure changes in the dynamic of chronicization.

Taken together, these results indicate that education is relevant for people with chronic low back pain. Additional investigations are necessary to measure the long-term effects on physical activity of these educations. Moreover, from literature reviews show positive effects for both educations (although that of school back education is more uncertain [10, 24]), we can suggest that some patients could better benefit from the back school while others would benefit more from pain neuroscience education depending on their profile (age, pain intensity, kinesiophobia, self-efficacy, occupational performance, etc.) and on their attitudes (levels of physical activity).

### Objectives

The main objective of the study is to evaluate the effectiveness of a pain neuroscience education on physical activity three months after the intervention compared to a school back education in patients with chronic low back pain attending a multidisciplinary rehabilitation program. The main measure is the average number of steps taken by participants over a week at home. This variable is at the heart of our study because we believe that it is on the one hand a criterion for judging the functional benefit of an education program, and on the other hand, in the extension of recent work on the chronic low back pain, a main criterion to measure behavioral changes in chronic low back pain essential to modify the dynamics of chronicization by physical activity.

The secondary objectives are grouped in three families. The first one concerns the comparison of the effectiveness of the two education programs at three months and one year, on other variables measuring physical activity, occupational performance, pain intensity, central sensitization, psychological variables specific to chronic pain (catastrophizing and kinesiophobia) and quality of life. The second one concerns the comparison of the changes of these variables over time, between the two groups. The third one concerns exploratory analyses to determine the predictors of the maintenance of physical activity for each of the programs in order to determine the success factors of the programs.

## METHODS AND ANALYSIS

### Trial design

This single-center study is prospective, open-label, controlled, randomized, superiority, with two parallel arms: an experimental “pain neuroscience education” group and a control “school back education” group. The duration of the inclusions will be thirty-four months and the duration of participation for each patient will be fourteen months. After his request for hospitalization, the investigation team will contact the patient to inform him and present the study, then, if the patient agrees, check the inclusion and exclusion criteria. If the patient is eligible and agrees, the patient will receive the study information letter (V0). A meeting will then be arranged. During this, the patient will then benefit from a pre-admission medical consultation to which is added a visit with one of the study’s investigators. This visit (V1) will make it possible to recheck the eligibility criteria, provide the necessary information to the patient and obtain the patient’s consent. The purpose of the visit will be to place an actigraph on the patient, which he will keep for seven days. Fifteen days to one month after this visit, the patient will be admitted to the establishment (V2). The hospitalization will be carried out as a part-time hospitalization (patient is discharged at weekends) and will last four weeks. During hospitalization, the patient will benefit from a multidisciplinary rehabilitation program and ten education sessions (pain neuroscience education or back school). The patient will then be seen again three months after discharge (V3) for a new pose of the actigraph that will be worn for a week. Finally, a last visit will be carried out one year before the release (V4) for a final installation of the actigraph (cf. figure 1).

**Figure 1:**
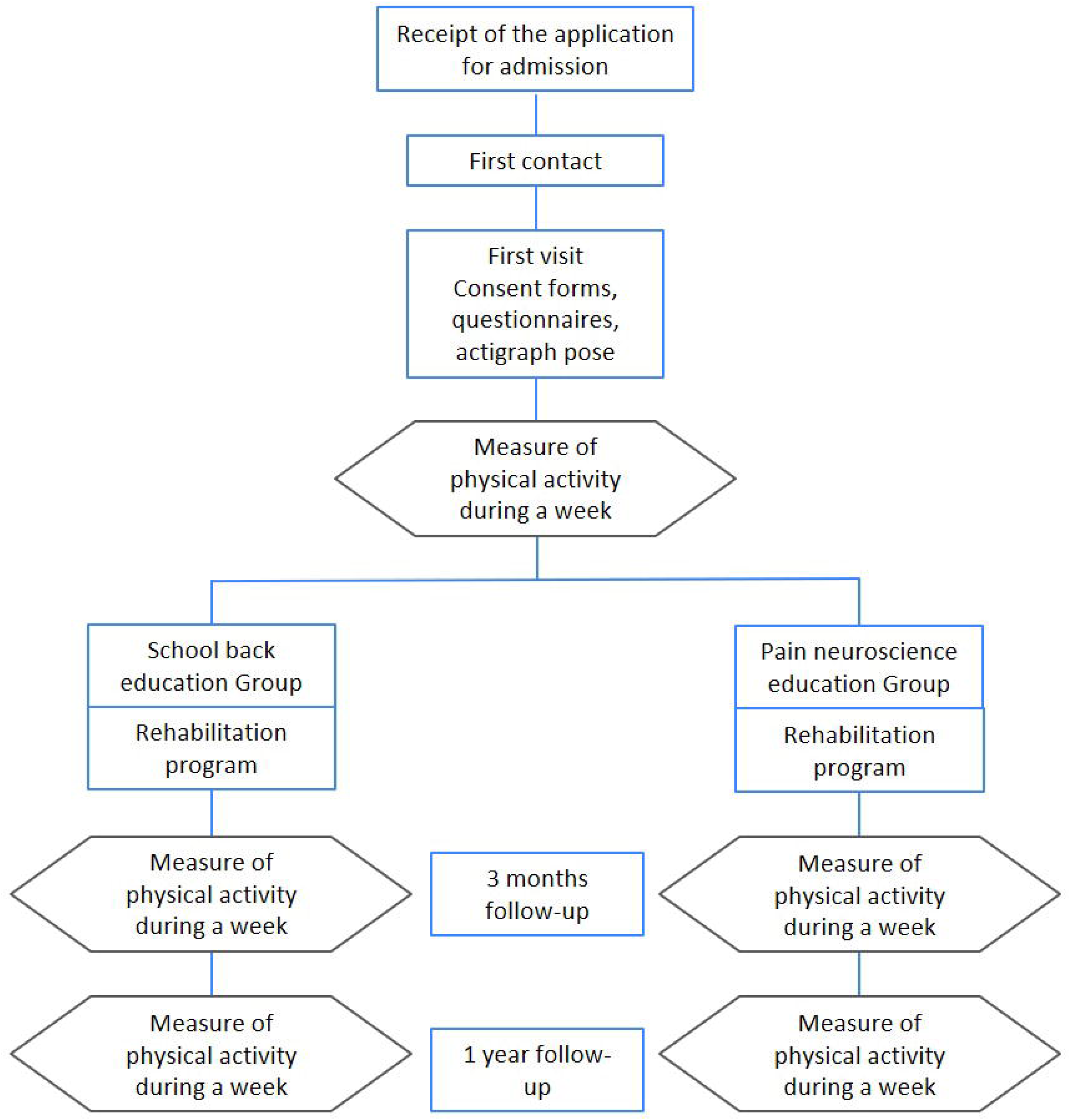
flow chart of the study design

### Participants

Eighty-two participants with chronic low back pain will take part in the study. All participants will benefit from specific education (pain neuroscience education vs back school) coupled with a multidisciplinary rehabilitation program. The study will be conducted in a French hospital center (Lamalou-les-bains, Occitanie).

#### Inclusion criteria

1. Patient over 18 years old.
2. Common low back pain according to HAS 2019^2^ criteria [25].
3. Chronic low back pain (> 3 months).
4. Start Back Screening Tool score > 3 (presence of psychosocial factors associated with a medium or high risk or chronic low back pain).
5. Part-time hospitalization at the hospital center for a multidisciplinary rehabilitation program.

#### Exclusion criteria

1. Subject with a gait-limiting comorbidity (e.g. central neurological disorder).
2. Subject with current psychiatric or cognitive comorbidity that does not allow education programs to be carried out.
3. Surgical intervention less than three months ago.
4. Other specific treatment for low back pain planned during the 3 months of follow-up (surgery, infiltration).
5. Patient participating in another clinical trial related to low back pain.
6. Subject not understanding the French language.
7. Pregnant, parturient or breast-feeding women.
8. Subject having a measure of legal protection (tutorship, curatorship).
9. Subject under safeguard of justice.
10. Subject who did not sign the informed consent form.
11. Subject not affiliated to a social security scheme or not a beneficiary of such a scheme.

### Randomization

The randomization will be carried out after obtaining the level of activity, measured by the actigraph during the week following V1. It will be balanced and centralized using the Ennov Clinical® software. A minimization algorithm will balance the level of activity between groups. We will use the levels of activity defined by Lotzke and colleagues [26], that is: sedentary (< 5000 steps by day), low activity (between 5000 and 7499 steps by day), active (between 7500 and 10000), and very active (>10000 steps by day).

### Blinding

Due to the type of intervention, participants and staff will know the randomization arm.

### Risk of contamination bias

In order to avoid that the participants know the content of the educational program of the other group, they will carry out their hospitalization in two different sites of the same hospital center. This is important not only for the sake of the power of the study, but also because mixing these two educational programs may decrease their efficacy. However, it is important to note that apart from the educational program, the content of the rehabilitation program is strictly similar between the two sites. Moreover, a close follow-up of patients during the rehabilitation will allow to assess precisely potential protocol deviations.

### Interventions

Both groups will benefit from ten education sessions and a multidisciplinary rehabilitation program. The educational sessions will be carried out in different dedicated rooms (in the respective places of care). The groups will consist of three to six participants. Some people may not be part of the study but they will all have a problem of chronic low back pain.

#### Pain Neuroscience Education Group

The educational sessions of the pain neuroscience education group are based on the work of Moseley and colleagues [14,17]. The target concepts are presented in table 1. The heart of education consists in considering pain not in the head, nor in the brain or only in the body or in connection with our thoughts and our emotions, but through all of these aspects (i.e. biopsychosocial model). Chronic pain is multifactorial and cannot be explained solely through x-rays, for example. This education aims, during the sessions, to modify misconceptions about pain and change beliefs (i.e. catastrophism). Moreover, the objective of this understanding of pain is to allow the participant to make changes in their attitudes, actions and thoughts when pain is perceived, or more generally to act in a more appropriate way in their environment.

**Table 1:**
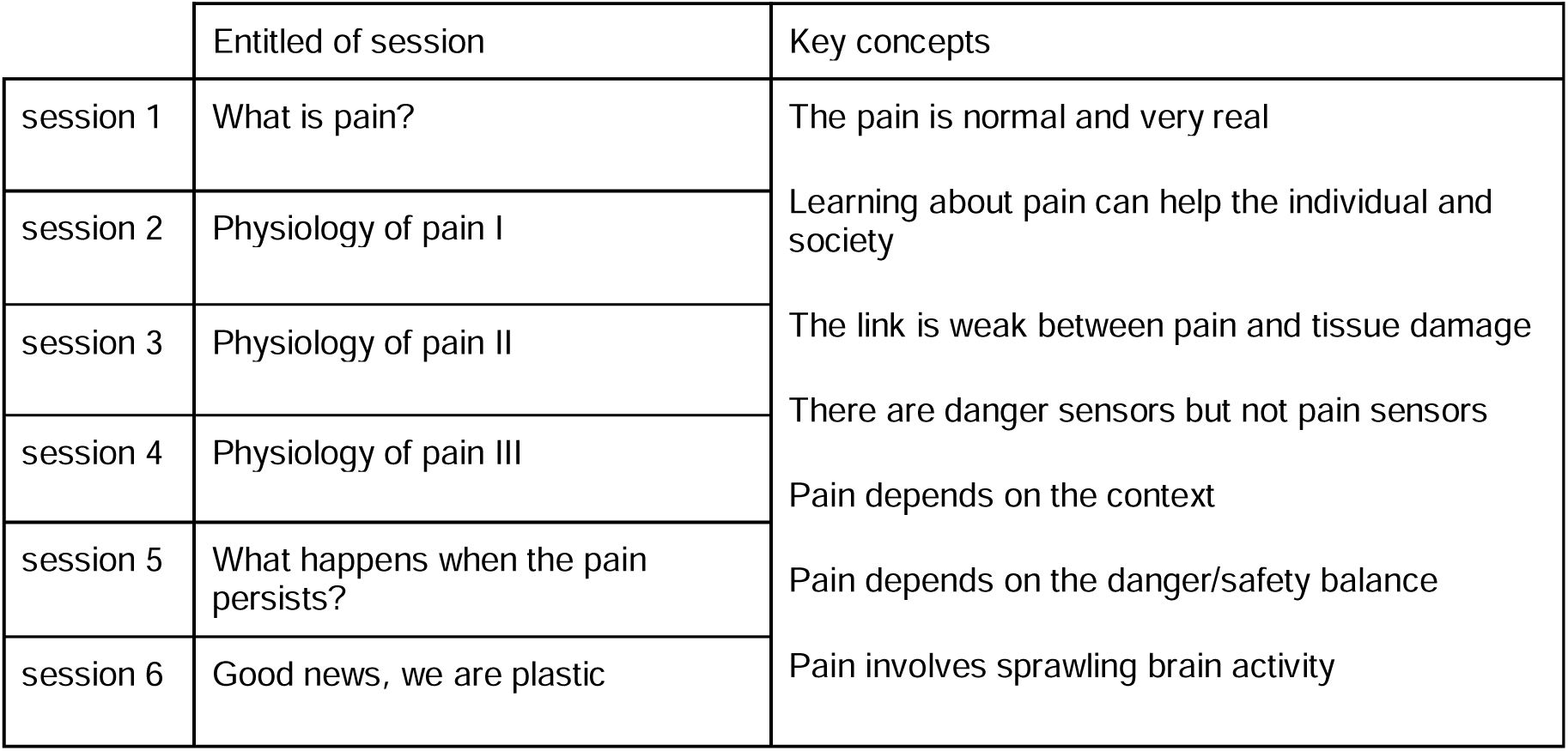
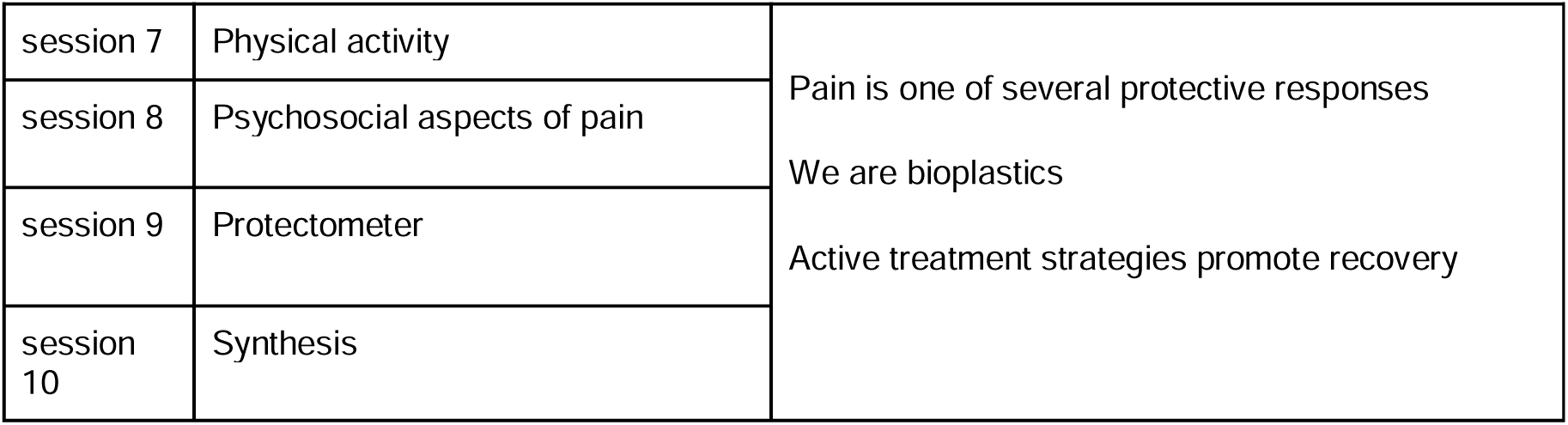
pain neuroscience education sessions.

#### School back education

Back school sessions are based on the conservative and preventive idea following back pain, in the spirit of Forsell’s work [8]. This back school consists of promoting prophylactic gestures during daily activities (e.g. bending down to lift a load). The program will consist of presenting the anatomical bases of the back and anatomo-physiology related to back pain [9, 27]. The table 2 presents the different elements that will be covered during the back school.

**Table 2:** school back education sessions care.

|  | Entitled of session | Key concepts |
| --- | --- | --- |
| session 1 | Anatomo-pathological reminders I | The structure of the spine, disc<br>Curvatures<br>Spinal pathologies<br>Specific mattresses and pillows<br>The positions<br>turn around, get up, lie down<br>The different sitting positions<br>The position in front of a computer<br>Principles and techniques of charging<br>The different positions for catching a load<br>Move with a load<br>Demonstration or Realization of activities taking into account the principles of spinal economy<br>The positions to be preferred according to the type of pain and the partner suffering from chronic low back pain |
| session 2 | Anatomo-pathological reminders II |  |
| session 3 | Lying position |  |
| session 4 | Sitting |  |
| session 5 | Gestures and postures |  |
| session 6 | Domestic activities I |  |
| session 7 | Domestic activities II |  |
| session 8 | Hobbies |  |
| session 9 | Car driving |  |
| session 10 | Sexuality |  |

#### Rehabilitation program

The four-week multi-professional rehabilitation program is part of the patient’s usual

This program will include daily, five days a week:

- Collective sessions of 30 minutes of adapted physical activity (APA) (muscle awakening, aerobic exercises, muscle strengthening, relaxation and muscle stretching).
- One or two 30-minute individual physiotherapy sessions (assessment, targeted strengthening exercises, motor control, self-exercise training, etc.).
- A 30-minute collective balneotherapy session (aquagym).
- A 20 to 30 minute session of physiotherapy (cryotherapy, thermotherapy, analgesic electrotherapy, etc.).

In addition, the patient will benefit from group relaxation sessions, an individual dietary consultation during his stay (assessment and dietary recommendation), and one or more psychological consultations for evaluative or therapeutic purposes.

Each patient will be given a form allowing them to check off the activities they carry out daily, this form will be returned to one of the investigators at the end of the stay. It will make it possible to check the volume of rehabilitation carried out in each group.

### Outcomes measures

#### Primary outcome

The primary outcome uses the actimetry medical device ActiGraph wGT3X-BT and the associated software, “ActivLife”, whose reliability and validity of the measurements of the number of steps and activity in an ecological situation are robust [28, 29]. The ActiGraph wGT3X-BT captures and records high-resolution raw acceleration data, which is converted into a variety of objective activity measurements using validated algorithms. As recommended by Migueles and colleagues [30], the device will be worn at the right hip, USB port cover up, using an elastic waistband, for seven days, during awake time, excluding wet activities. The sampling frequency will be 30 Hz, we will use time windows of 1s for the categorization of activities (e.g. such as standing, sitting or lying down) [31].

The measurements will be exported in Comma-separated values (CSV) format from the “Activlife” software, the file will be anonymized according to the procedure provided for in the protocol, and stored on the hospital server.

#### Secondary outcomes

- Intensity of pain will be measured by the numerical rating scale (NRS) [25] (value between 0 and 10). The value indicated by the participant will represent the raw data used. Data will be collected during the enrolment, at three months and one year.
- Central nervous system pain sensitization will be evaluated by the Central Sensitization Questionnaire [32] at the enrollment and at three months and one year.
- Psychological variables will be explored by the catastrophizing scale [33], the kinesiophobia scale [34] and the Hospital Anxiety and Depression Scale (HADS [35]) at the enrollment and at three months and one year.
- Physical activity and occupation will be measured by the global questionnaire on the practice of physical activities (GPAQ [36]) and the Canadian Measure of Occupational Performance (COPM [37]) during the first week of rehabilitation and at three months.
- Quality of life will be evaluated by the SF-36 [38]. Data will be collected during the enrolment, at three months and one year.

#### Additional data

These data will be collected at the entrance to the hospital center:

- The questionnaire on the ability to change in the face of pain [39]. This self-questionnaire measures the degree of aptitude for change.
- The Chronic Pain Acceptance Questionnaire [40] indicates psychological flexibility in the face of pain.
- The Agency scale [41] measures the feeling that the person has to determine himself as the actor of his own actions.
- Start Back Screening Tool to define the degree of chronicization [42].
- Sociodemographic data: age, sex, socioeconomic level (profession, level of study), marital status.
- Clinical and medical data: height, weight, BMI, main diagnosed spinal pathology, main history, anteriority of the pathology, consumption of pain-related treatments (analgesics, anti-inflammatories)

Table 3 provides an overview of measurements by steps of the study.

**Table 3:**
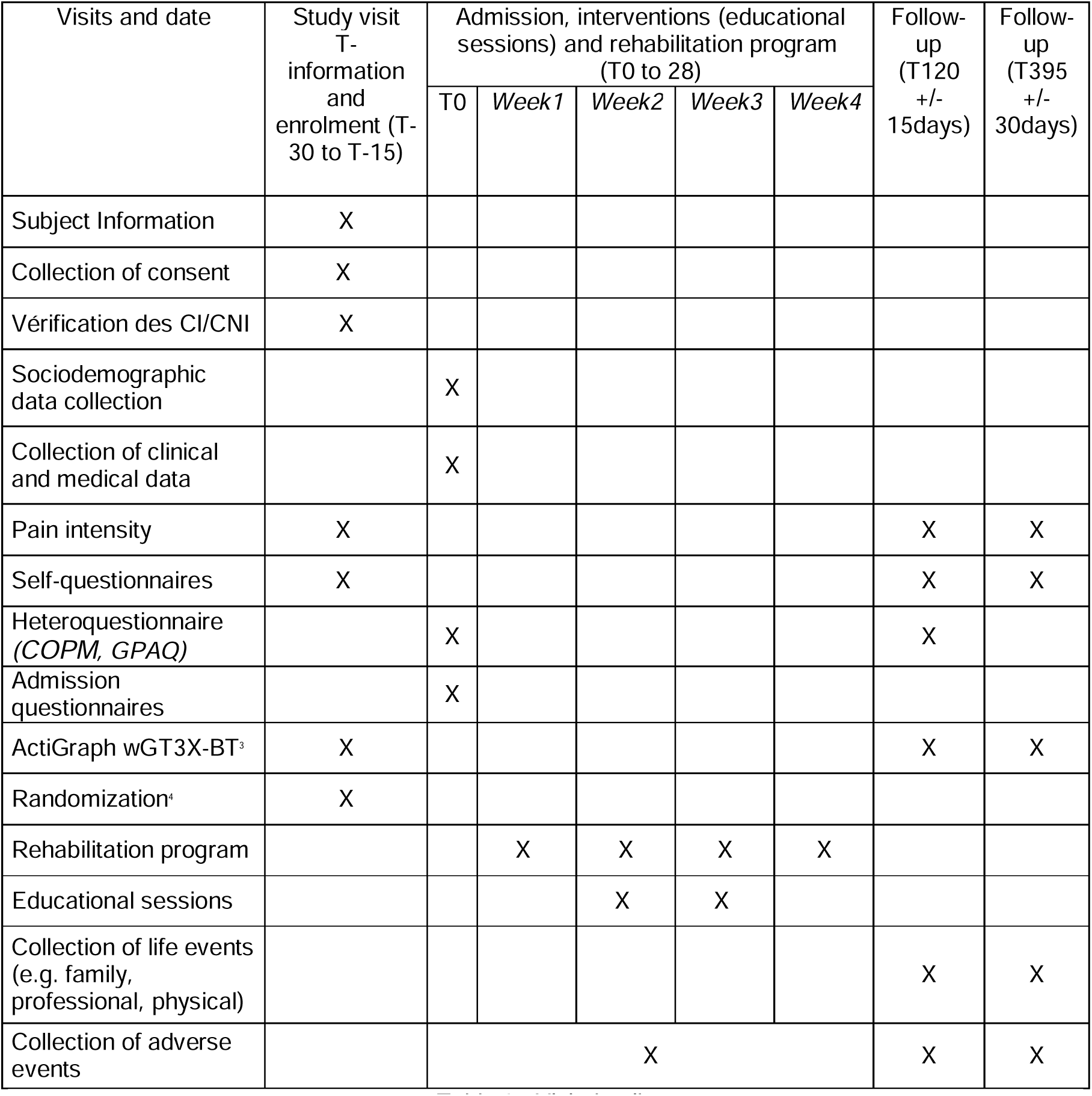
Visit details.

| Visits and date | Study visit<br>T-information<br>and<br>enrolment (T-30 to T-15) | Admission, interventions (educational<br>sessions) and rehabilitation program<br>(T0 to 28) |  |  |  |  | Follow-up<br>(T120<br>+/-<br>15days) | Follow-up<br>(T395<br>+/-<br>30days) |
| --- | --- | --- | --- | --- | --- | --- | --- | --- |
|  |  | T0 | Week1 | Week2 | Week3 | Week4 |  |  |
| Subject Information | X |  |  |  |  |  |  |  |
| Collection of consent | X |  |  |  |  |  |  |  |
| Vérification des CI/CNI | X |  |  |  |  |  |  |  |
| Sociodemographic<br>data collection |  | X |  |  |  |  |  |  |
| Collection of clinical<br>and medical data |  | X |  |  |  |  |  |  |
| Pain intensity | X |  |  |  |  |  | X | X |
| Self-questionnaires | X |  |  |  |  |  | X | X |
| Heteroquestionnaire<br>(COPM, GPAQ) |  | X |  |  |  |  | X |  |
| Admission<br>questionnaires |  | X |  |  |  |  |  |  |
| ActiGraph wGT3X-BT<sup>a</sup> | X |  |  |  |  |  | X | X |
| Randomization<sup>a</sup> | X |  |  |  |  |  |  |  |
| Rehabilitation program |  |  | X | X | X | X |  |  |
| Educational sessions |  |  |  | X | X |  |  |  |
| Collection of life events<br>(e.g. family,<br>professional, physical) |  |  |  |  |  |  | X | X |
| Collection of adverse<br>events |  | X |  |  |  |  | X | X |

The hetero questionnaires will be carried out by trained professionals and experts in the field (e.g. occupational therapist for COPM).

### Sample size calculation

In their cross-sectional study of 118 patients with chronic low back pain, Lotzke and colleagues [26] measured that 56% of their sample took less than 7,500 steps per day and 16% took only 5,000. The World Health Organization recommends 10,000 steps per day and the Haute Autorité de Santé in France, in its physical activity promotion guide recommends a gradual increase from 1,000 to 3,000 daily steps in order to adapt to individual abilities. For our study, we chose a difference between the two groups of 2000 daily steps, which corresponds to the average recommendation of the HAS [25]. The normative value of the primary endpoint is 8609 daily steps (standard deviation 2625) [43]. To show a significant difference between the groups at alpha risk of 5%, and with a power of 90%, it is necessary to analyze 37 patients per group. Taking into account a rate of loss of follow-up of 10%, it will be necessary to recruit 41 patients per group, i.e. 82 patients in total.

In order to take into account patients who have not started the multidisciplinary rehabilitation program, we plan to include up to a maximum of 100 subjects; inclusions will be stopped as soon as 41 patients per group are obtained.

### Statistical analysis

The significance threshold is set at 5% on the basis of a two-sided test. All analyzes will be carried out under SAS.

#### Analysis population

The included population (IP) is the entire population included in the study. The intent-to-treat (ITT) population includes all randomized patients in whom the primary endpoint will be effectively measured. The per-protocol population includes all patients who have been randomized and who have participated in at least 90% of the education sessions. The safety population is all included and randomized patients who have completed at least one education session.

#### Descriptive analysis

A simple descriptive analysis will be carried out on the entire ITT population and then on the two randomization groups. This analysis will cover all the data from the study. The continuous variables will be described by mean, standard deviation, median and quartiles, and the qualitative variables by their counts and percentages. A flow diagram will be produced from the included population.

#### Analysis of the primary and secondary outcomes

The analysis will be carried out on the ITT population. We will first check the normality of the distribution of the results in the two groups. Then we will carry out, in case of normality of the distributions, a parametric test of the reduced deviation or non-parametric Mann-Whitney test in case of absence of normality. If the p-value of the test is below the 5% threshold, then the program followed by the group with the highest average will be considered superior. The difference between the means of the two groups will be presented with its 95% confidence interval. The same procedure as that of the primary outcome will be carried out for the secondary outcomes. To control the alpha risk within each of the first two families of secondary outcomes, a Hochberg procedure will be applied.

#### Exploratory analysis

In addition to the data directly calculated by the Activlife software (type of activity and time spent on each activity, number of steps, energy expended), the raw accelerometric data will be analyzed. Different movement markers (acceleration, jerk, orientation) will be calculated at each instant, which will make it possible to finely characterize the activity of the subjects. In addition, the factors potentially associated with the practice of physical activity at 3 months and at one year will also be included in this analysis (i.e. pain intensity, catastrophizing, central sensitization, kinesiophobia, number of steps before the rehabilitation program, capacity for change, acceptance of chronic pain, agency, anxiety and depression, socio-demographic, clinical and medical data). The association between each of these data (accelerometric and other) measured at baseline, and the number of steps at 3 months and at one year, will first be evaluated in univariate, on the entire intention-to-treat population, by a potentially predictive variable model. Secondarily, the interaction between the group and the variable will be included in the model to look for a difference in the effect of pain neuroscience education, compared to back school, depending on this variable. The eight variables most associated with the number of steps will then be introduced into a multivariate model.

#### Management of changes to the initial statistical plan

Any modification to the initial statistical analysis plan will be discussed between the investigator, the methodologist, and the sponsor, and will be the subject of a request for substantial modification to the Committee for the Protection of Persons.

#### Management of missing data

Patients wishing to abandon the program before the end of the 4 weeks can complete the evaluation of the primary endpoint (V3), if they agree. The values of the endpoints obtained at the end of the study will be used for the analyses. If the rate of missing data is greater than 5% for a judgment criterion, a sensitivity analysis by multiple imputation will be carried out on this judgment criterion.

### Patient and public involvement

There was no patient or public participation in the design of this study (definition of the research question or outcome measures). Patients will not be asked to participate in the analysis and interpretation of the results of this study.

## Data Availability

All data produced in the present study are available upon reasonable request to the authors

## DATA AVAILABILITY

The data that support the findings of this study are available from the corresponding author, upon reasonable request.

## DATA SECURITY AND HANDLING

The study data will be entered into an electronic information notebook developed using the ENNOV CLINICAL® software, which allows real-time data quality control. The self-questionnaires will be completed online directly by the patient via the ENNOV CLINICAL® software. The connection to the self-questionnaires is done by a password and a unique identifier specific to each “patient” user. All “patient” user data is stored encrypted in the database (AES 128 bit encryption). The patient’s personal information that may be stored (surname, first name, email) is only visible when the patient is created. So, the patient’s identity will not be disclosed.

The data will be accessible to the principal investigator, the project leader and the methodologists associated with the project.

## ETHICS AND DISSEMINATION

The participant’s signed informed consent will be collected during the inclusion visit and before any study-specific procedures are performed. A copy of the signed consent will be given to the patient.

A favorable opinion was given by the Committee for Personal Protection of Ile de France VII on June 22, 2023 (National number: 2023-A00346-39). The study was previously registered with the National Agency for the Safety of Medicines and Health Products (IDRCB : 2023-A00346-39).

The results collected from this protocol will be presented at national or international conferences. The results will be published in peer-reviewed journals aimed at physicians, medical auxiliaries (physiotherapists, occupational therapists), psychologists and in the field of therapeutic education. However, the promoter will send the Committee for the Protection of Persons the results of the research in the form of a summary of the final report within one year after the end of the research.

Any written or oral communication of research results must receive the prior agreement of the coordinating investigator.

## AUTHOR CONTRIBUTIONS

Gaël Le Perf and Guillaume Thébault developed the study idea and elaborated the scientific aspects of the study. Gaël Le Perf, Guillaume Thébault, and Claire Duflos elaborated the methodological aspects of the study. Gaël Le Perf submitted the project for financing. The authors jointly developed the final protocol. Gaël Le Perf is the principal investigator of the study. Guillaume Thébault is a project leader and associate investigator. Claire Duflos is the study methodologist. Guillaume Thébault formatted the article for publication.

## FUNDING STATEMENT

This study is supported by the Ministry of Solidarity and Health under a Hospital Program for Nursing and Paramedical Research (Grant Number: PHRIP-20-0259) and by the National Council of the Order of Physiotherapist (CNOMK) (Grant Number: 2022-03).

## COMPETING INTERESTS STATEMENT

None declared.

## SPONSOR

University Hospital Center, Montpellier is the promoter of the study. The Centre Hospitalier Paul Coste Floret is a sponsor. Gaël Le Perf is the principal investigator of the study.

## ACKNOWLEDGEMENTS

We thank Sylvie Grandemange, of the Research and Innovation Department of the CHU Montpellier, Claire Belloc, Denis Mottet and Philippe Mugnier for their advice and support. We would also like to thank the entire hospital team who invested in the realization of this study.

## Footnotes

1 In french: “le bon traitement, c’est le mouvement”.

2 “Common low back pain is defined by pain located between the thoracolumbar hinge and the lower gluteal fold (AE) [which may] be associated with radiculalgia corresponding to pain in one or both lower limbs at the level of one or more dermatomes (AE) which do not show warning signs (see “red flags”)” (HAS, 2019).

## Notes

### Competing Interest Statement

The authors have declared no competing interest.

### Clinical Trial

NCT05840302

### Funding Statement

This study is supported by the Ministry of Solidarity and Health as part of a hospital nursing and paramedical research program (grant number: PHRIP-20-0259) and by the National Council of the Order of Physiotherapists (CNOMK) (grant number: 2022-03).

### Author Declarations

The Ile de France VII Personal Protection Committee gave its agreement on June 22, 2023 for this work (National number: 2023-A00346-39) The National Agency for the Safety of Medicines and Health Products approved this work (CRDI: 2023-A00346-39)

